# Screening Renin-Aldosterone Profiles and Long-term Cardiovascular Risk After Primary Aldosteronism Evaluation

**DOI:** 10.64898/2026.11.09.26367682

**Authors:** Heng-Chih Pan, Kuang-Hung Chen, Cheng-Hsuan Tsai, Yen-Hung Lin, Cheng-Yi Wang, Vin-Cent Wu

## Abstract

**Background:** Aldosterone-to-renin ratio (ARR) and plasma renin activity (PRA) are measured at the screening stage of primary aldosteronism (PA) evaluation, but their prognostic relevance after confirmatory classification and subsequent treatment remains uncertain.

**Methods:** We linked the seven-hospital Taiwan Primary Aldosteronism Investigation cohort (1991– 2023) with nationwide health insurance and mortality data through June 2025. Patients were classified as confirmatory-test-defined essential hypertension (EH), PA without adrenalectomy, or PA with subsequent adrenalectomy. Covariate-adjusted Cox models evaluated screening logARR, logPRA, and logPAC separately in relation to incident major adverse cardiovascular events (MACE; myocardial infarction, stroke, heart failure hospitalization, or cardiovascular death).

**Results:** Among 3,178 participants (EH, 1,433; PA without adrenalectomy, 834; PA with adrenalectomy, 911), 441 (13.9%) experienced MACE during a median 7.9-year follow-up. Adjusted hazard ratios (95% CIs) per 1-unit increase in logARR were 1.09 (1.00–1.19), 1.14 (1.03–1.26), and 0.95 (0.87–1.05), respectively; corresponding logPRA estimates were 0.91 (0.83–0.99), 0.87 (0.79– 0.97), and 1.04 (0.93–1.16). LogPAC showed no consistent associations. Primary biomarker-by-phenotype interactions were not significant for logARR (P=0.057) or logPRA (P=0.069); time-varying adrenalectomy analyses were supportive but exploratory.

**Conclusions:** Higher screening ARR and lower PRA were associated with subsequent MACE in confirmatory-test-defined EH and PA without adrenalectomy, but not clearly in PA with subsequent adrenalectomy.

## Introduction

Primary aldosteronism (PA) is a common, underdiagnosed, and treatable cause of hypertension associated with excess cardiovascular and kidney risk beyond blood pressure elevation^1–3^. At the screening stage, paired aldosterone and renin measurements guide initial PA evaluation^1,4^. The aldosterone-to-renin ratio (ARR) reflects aldosterone relative to renin suppression, whereas plasma renin activity (PRA) and plasma aldosterone concentration (PAC) are its components^5,6^; these measures are related but not interchangeable^5–7^.

Renin-independent aldosterone physiology spans subclinical or low-renin states to overt PA^6,8–10^. Higher ARR and lower renin have been associated with hypertension, cardiovascular events, and kidney function decline, whereas aldosterone alone has shown less consistent associations^8,11–14^.

However, prior population-based studies did not distinguish confirmatory-test-defined essential hypertension (EH) from PA or account for subsequent PA treatment pathways^12–14^. Conversely, clinical PA studies have not focused on the long-term prognostic significance of screening ARR and PRA^15–18^. Thus, whether screening renin–aldosterone profiles provide prognostic information beyond diagnostic classification in the real-world PA evaluation pathway remains uncertain. Using a large hypertension cohort linked to nationwide health insurance data, we evaluated screening ARR, PRA, and PAC in parallel and examined their associations with incident major adverse cardiovascular events (MACE) among confirmatory-test-defined EH, PA without adrenalectomy, or PA with adrenalectomy. We further examined whether higher ARR and lower PRA showed concordant risk patterns after diagnostic classification and subsequent treatment-pathway stratification.

## Methods

### Study Design and Data Source

We conducted an observational cohort study using the multicenter Taiwan Primary Aldosteronism Investigation (TAIPAI) database linked with Taiwan’s National Health Insurance Research Database (NHIRD). TAIPAI has enrolled patients undergoing ARR screening for possible PA since June 1, 1991, across 7 hospitals in Taiwan^19–21^. The index date was the screening-stage visit before confirmatory testing, when PRA and PAC were measured and ARR was calculated. Index dates after June 30, 2023, were excluded; outcomes were available through June 30, 2025. The National Taiwan University Hospital institutional review board approved the study (200611031R), and all participants provided written informed consent.

### Study Population and Clinical Phenotypes

Adults were eligible if they underwent PA evaluation with baseline PRA and PAC measured before confirmatory testing. Exclusions were a missing or post-June 30, 2023 index date, unavailable baseline PRA or PAC, adrenalectomy before index, or qualifying MACE before index. PA was confirmed by autonomous aldosterone production according to the TAIPAI framework, including validated post-captopril or post-losartan ARR criteria^19,20,22^.

Patients were classified as (1) EH, without confirmatory evidence of PA; (2) PA without adrenalectomy; or (3) PA with subsequent adrenalectomy (**Figure 1**). Because adrenalectomy occurred after PA evaluation and was not randomized, these analyses were interpreted as treatment-pathway-stratified prognostic associations rather than causal effects of surgery^23,24^.

**Figure 1.**
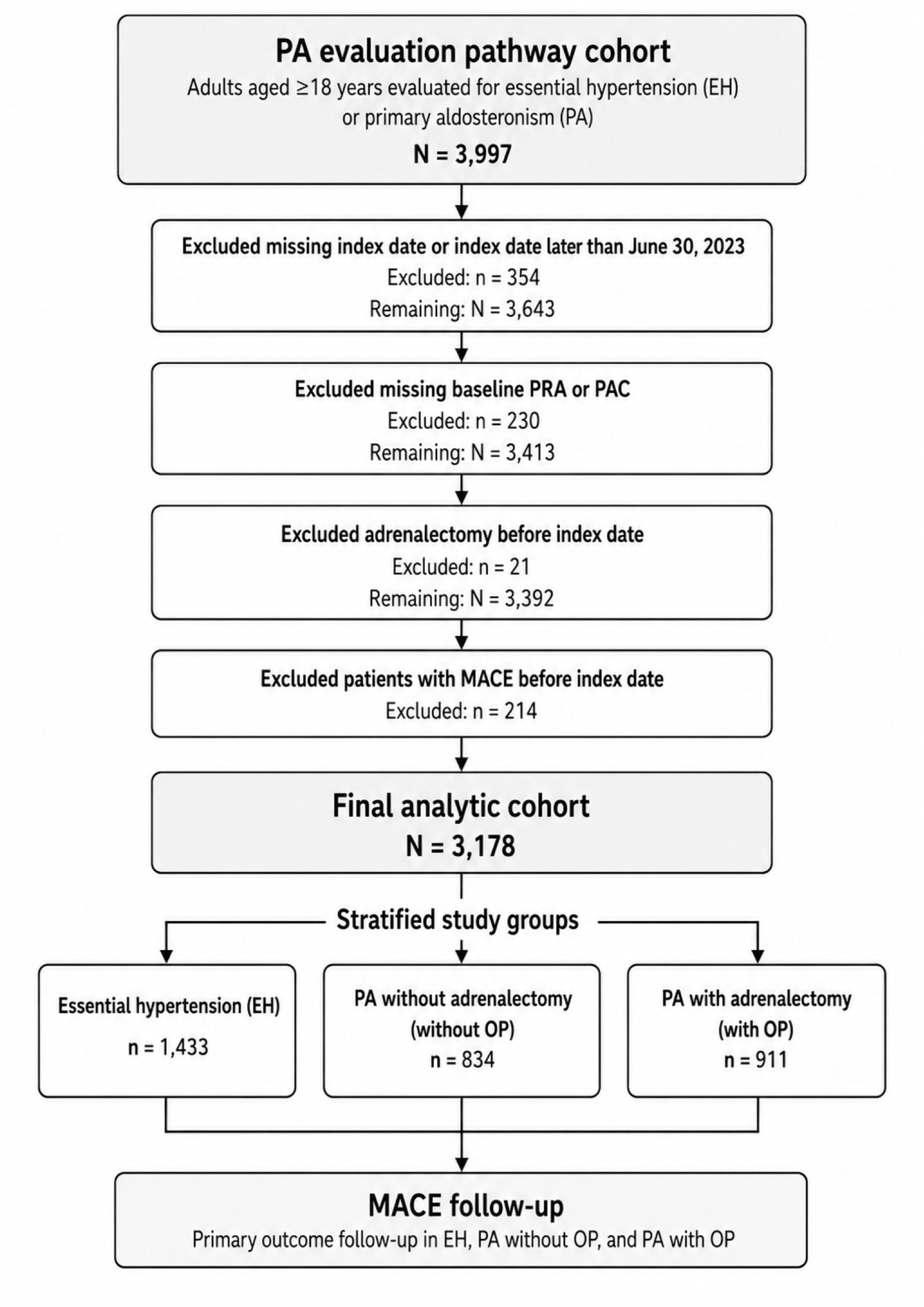
Study flow and PA evaluation pathway. * The index date was the baseline PA screening visit.

### Identification of PA and Biomarker Measurement

Other causes of secondary hypertension were excluded according to TAIPAI procedures^20,25^. Potentially interfering antihypertensive medications were withheld for at least 21 days when clinically appropriate; doxazosin and/or diltiazem were used when needed, and usual sodium intake was maintained and assessed by urinary testing^1,20^. PA was diagnosed according to TAIPAI and Taiwan Society of Aldosteronism criteria^19,20,22,25^; detailed confirmation and subtype procedures are provided in the Supplementary Methods.

PAC and PRA were measured by radioimmunoassay (ALDO-RIACT, Cisbio Bioassays; Ang I RIA, Beckman Coulter). The aldosterone detection limit was 10.0 pg/mL; upright reference ranges were 70–350 pg/mL for PAC and 2.63 ± 1.32 ng/mL/h for PRA. PRA intra-assay and interassay coefficients of variation were 1.9% and 4.5%.

### Exposure Assessment

Screening ARR and PRA were defined as measurements obtained at the initial PA evaluation after a medication washout period for antihypertensive agents that could interfere with the renin– aldosterone system, whenever clinically feasible. ARR, PRA, and PAC measured at the screening/index visit were natural-log transformed because of right-skewed distributions and analyzed in separate models with identical covariate adjustment. No data-driven ARR or PRA cutoffs were used because thresholds vary with assay and clinical conditions^1,5,7^.

### Outcomes

Outcomes were ascertained by linking TAIPAI to Taiwan’s NHIRD, which covers approximately 99% of the population^26^. Baseline clinical and laboratory data came from TAIPAI; claims and mortality data were linked using encrypted identifiers and national data services^27^. Cause-specific mortality was obtained from death certificates.

The primary outcome was MACE, defined as incident myocardial infarction, stroke, heart failure hospitalization, or cardiovascular death. Baseline heart failure in **Table 1** reflected comorbidity at study entry; patients with a qualifying heart failure hospitalization before the index date were excluded, and the heart failure component of MACE required a hospitalization after the index date. Secondary outcomes were individual MACE components and all-cause mortality. Claims-based cardiovascular definitions have been previously validated^28,29^, and stroke ascertainment used diagnostic codes with neuroimaging information^30^. Detailed ICD definitions are provided in **eTable 1**. Follow-up began at index and continued until first MACE, noncardiovascular death, loss to follow-up, or study end.

**Table 1.** Baseline characteristics of the final analytic cohort by clinical phenotype.

| Variable | Total<br>(N = 3,178) | EH<br>(N = 1,433) | PA without OP<br>(N = 834) | PA with OP<br>(N = 911) |
|---|---|---|---|---|
| Age, years | 52.1 ± 13.5 | 50.8 ± 14.9 | 55.6 ± 12.6 | 50.9 ± 11.4 |
| Age ≥65 years, n (%) | 577 (18.2) | 273 (19.1) | 201 (24.1) | 103 (11.3) |
| Female sex, n (%) | 1,668 (52.5) | 717 (50.0) | 458 (54.9) | 493 (54.1) |
| Body mass index, kg/m² | 25.5 ± 4.1 | 25.4 ± 4.1 | 25.4 ± 3.9 | 25.6 ± 4.2 |
| Systolic BP, mmHg | 148.6 ± 21.5 | 144.5 ± 20.1 | 149.6 ± 21.9 | 154.1 ± 22.2 |
| Diastolic BP, mmHg | 88.8 ± 14.1 | 86.1 ± 13.4 | 89.1 ± 14.1 | 92.6 ± 14.5 |
| **Laboratory measures** |  |  |  |  |
| Potassium, mmol/L | 3.8 ± 0.5 | 4.1 ± 0.3 | 3.8 ± 0.5 | 3.4 ± 0.6 |
| Creatinine, mg/dL | 0.9 ± 0.4 | 0.9 ± 0.4 | 0.9 ± 0.5 | 0.9 ± 0.4 |
| eGFR, mL/min/1.73 m² | 89.2 ± 25.6 | 89.5 ± 24.8 | 88.5 ± 26.4 | 89.3 ± 26.1 |
| Uric acid, mg/dL | 5.9 ± 1.6 | 6.0 ± 1.6 | 5.9 ± 1.6 | 5.8 ± 1.6 |
| HbA1c, % | 6.0 ± 1.0 | 5.9 ± 0.9 | 6.0 ± 1.0 | 6.0 ± 0.9 |
| LDL cholesterol, mg/dL | 113.6 ± 30.8 | 116.4 ± 31.1 | 112.8 ± 30.6 | 110.2 ± 30.2 |
| PRA, ng/mL/h | 0.6 [0.1, 1.7] | 1.4 [0.5, 3.7] | 0.4 [0.1, 0.9] | 0.3 [0.1, 0.7] |
| PAC, ng/dL | 34.1 [23.1, 51.7] | 30.4 [20.6, 45.4] | 34.8 [24.1, 51.5] | 40.0 [26.7, 63.2] |
| ARR, ng/dL per ng/mL/h | 53.6 [18.5, 220.2] | 20.5 [9.2, 56.7] | 97.8 [39.7, 304.7] | 156.1 [50.5, 520.0] |
| **Comorbidities** |  |  |  |  |
| Diabetes mellitus, n (%) | 469 (14.8) | 156 (10.9) | 153 (18.3) | 160 (17.6) |
| Hyperlipidemia, n (%) | 798 (25.1) | 372 (26.0) | 226 (27.1) | 200 (22.0) |
| Atrial fibrillation, n (%) | 69 (2.2) | 17 (1.2) | 29 (3.5) | 23 (2.5) |
| CAD*, n (%) | 233 (7.3) | 95 (6.6) | 71 (8.5) | 67 (7.4) |
| Baseline heart failure diagnosis*, n (%) | 19 (0.6) | 11 (0.8) | 4 (0.5) | 4 (0.4) |
| COPD, n (%) | 23 (0.7) | 12 (0.8) | 5 (0.6) | 6 (0.7) |
| **Medications** |  |  |  |  |
| ACE inhibitor, n (%) | 78 (2.5) | 27 (1.9) | 20 (2.4) | 31 (3.4) |
| ARB, n (%) | 1,202 (37.8) | 557 (38.9) | 302 (36.2) | 343 (37.7) |
| Beta-blocker, n (%) | 956 (30.1) | 316 (22.1) | 302 (36.2) | 338 (37.1) |
| Alpha-blocker, n (%) | 518 (16.3) | 159 (11.1) | 157 (18.8) | 202 (22.2) |
| CCB, n (%) | 1,846 (58.1) | 725 (50.6) | 494 (59.2) | 627 (68.8) |
| MRA, n (%) | 488 (15.4) | 63 (4.4) | 148 (17.7) | 277 (30.4) |
| Diuretics, n (%) | 260 (8.2) | 84 (5.9) | 91 (10.9) | 85 (9.3) |
| Vasodilator, n (%) | 146 (4.6) | 40 (2.8) | 45 (5.4) | 61 (6.7) |
| Number of antihypertensive drugs | 1.7 ± 1.2 | 1.3 ± 1.1 | 1.8 ± 1.2 | 2.1 ± 1.2 |

**Abbreviation:** ARR indicates aldosterone-to-renin ratio; CAD, coronary artery disease; CCB, calcium channel blocker; eGFR, estimated glomerular filtration rate; EH, essential hypertension; MRA, mineralocorticoid receptor antagonist; PA, primary aldosteronism; PAC, plasma aldosterone concentration; PRA, plasma renin activity.
\* Baseline coronary artery disease and heart failure represent recorded comorbidity status. Patients with a qualifying myocardial infarction or heart failure hospitalization before the index date were excluded; only qualifying events occurring after the index date contributed to MACE.
\* Medication use was assessed at the baseline screening visit and does not represent treatment initiated after PA confirmation or cumulative medication exposure during follow-up.

### Statistical Analysis

Baseline characteristics were summarized by clinical phenotype. Separate Cox models evaluated associations of logARR, logPRA, and logPAC with MACE within each phenotype, adjusting for age category, sex, body mass index, systolic blood pressure category, estimated glomerular filtration rate, and serum potassium; biomarker-by-phenotype interactions were tested separately. Hazard ratios and 95% confidence intervals were reported per 1-unit increase in each natural-log-transformed biomarker. Negative control analyses excluded participants with the corresponding pre-index outcome and modeled logARR and logPRA using the same covariates^31^.

Cox generalized additive models characterized dose-response patterns. Secondary analyses evaluated calendar-time effects, adrenalectomy timing, and negative control outcomes. All analyses were performed using Stata version 17 (StataCorp, College Station, TX, USA) and SAS version 9·4 (SAS Institute Inc., Cary, NC, USA). Two-sided *P*<.05 indicated statistical significance. Additional details are provided in the Supplementary Methods.

## Results

### Study Cohort and Baseline Characteristics

The final analytic cohort included 3,178 patients who underwent PA evaluation. Among them, 1,433 were classified as EH, 834 as PA without adrenalectomy, and 911 as PA with adrenalectomy (**Figure 1** and **Table 1)**.

Patients with PA had lower PRA and higher ARR than patients with EH. Median PRA was 0.6 [0.1–1.7] ng/mL/h in the overall cohort, 1.4 [0.5–3.7] in EH, 0.4 [0.1–0.9] in PA without adrenalectomy, and 0.3 [0.1–0.7] in PA with adrenalectomy. Median ARR was 53.6 [18.5–220.2] overall, 20.5 [9.2–56.7] in EH, 97.8 [39.7–304.7] in PA without adrenalectomy, and 156.1 [50.5–520.0] in PA with adrenalectomy. Median PAC was 34.1 [23.1–51.7], 30.4 [20.6–45.4], 34.8 [24.1–51.5], and 40.0 [26.7–63.2] ng/dL, respectively. Other baseline differences, including blood pressure, potassium, kidney function, comorbidities, and medication use, are summarized in **Table 1**.

### Clinical Outcomes

During a median follow-up of 7.9 years (IQR, 4.4–12.9), 441 patients (13.9%) experienced MACE. The overall MACE incidence rate was 16.3 per 1,000 person-years, with rates of 15.0, 21.4, and 14.3 per 1,000 person-years in EH, PA without adrenalectomy, and PA with adrenalectomy, respectively. Overall, myocardial infarction occurred in 107 patients (3.4%), heart failure hospitalization in 104 (3.3%), stroke in 260 (8.2%), cardiovascular death in 70 (2.2%), and all-cause death in 262 (8.2%) (**Table 2**).

**Table 2.** Incident clinical outcomes by clinical phenotype.

| Outcome | Total<br>(N = 3,178) | EH<br>(N = 1,433) | PA without OP<br>(N = 834) | PA with OP<br>(N = 911) |
|---|---|---|---|---|
| Follow-up time, median [IQR], years | 7.9 [4.4, 12.9] | 7.9 [4.4, 12.9] | 7.5 [3.4, 10.8] | 8.8 [5.2, 13.3] |
| MACE, n (%) | 441 (13.9) | 179 (12.5) | 137 (16.4) | 125 (13.7) |
| Myocardial infarction | 107 (3.4) | 47 (3.3) | 30 (3.6) | 30 (3.3) |
| Heart failure hospitalization | 104 (3.3) | 43 (3.0) | 33 (4.0) | 28 (3.1) |
| Cardiovascular death | 70 (2.2) | 27 (1.9) | 25 (3.0) | 18 (2.0) |
| Stroke | 260 (8.2) | 98 (6.8) | 84 (10.1) | 78 (8.6) |
| All-cause death | 262 (8.2) | 114 (8.0) | 81 (9.7) | 67 (7.4) |
| MACE incidence rate, per 1,000 person-years | 16.3 | 15.0 | 21.4 | 14.3 |

**Abbreviation:** MACE, major adverse cardiovascular events.
\*Individual participants could experience more than one MACE component; therefore, the sum of component events may exceed the number of patients with composite MACE.

### Screening Renin-Aldosterone Markers and Incident MACE (Table 3 and Figure 2)

Higher logARR was associated with higher MACE risk in EH (adjusted HR, 1.09; 95% CI, 1.00-1.19; P=0.046) and PA without adrenalectomy (adjusted HR, 1.14; 95% CI, 1.03-1.26; P=0.013), but not in PA with adrenalectomy (adjusted HR, 0.95; 95% CI, 0.87-1.05; P=0.348). The logARR-by-clinical-phenotype interaction did not reach conventional statistical significance (P for interaction=0.057).

**Figure 2.**
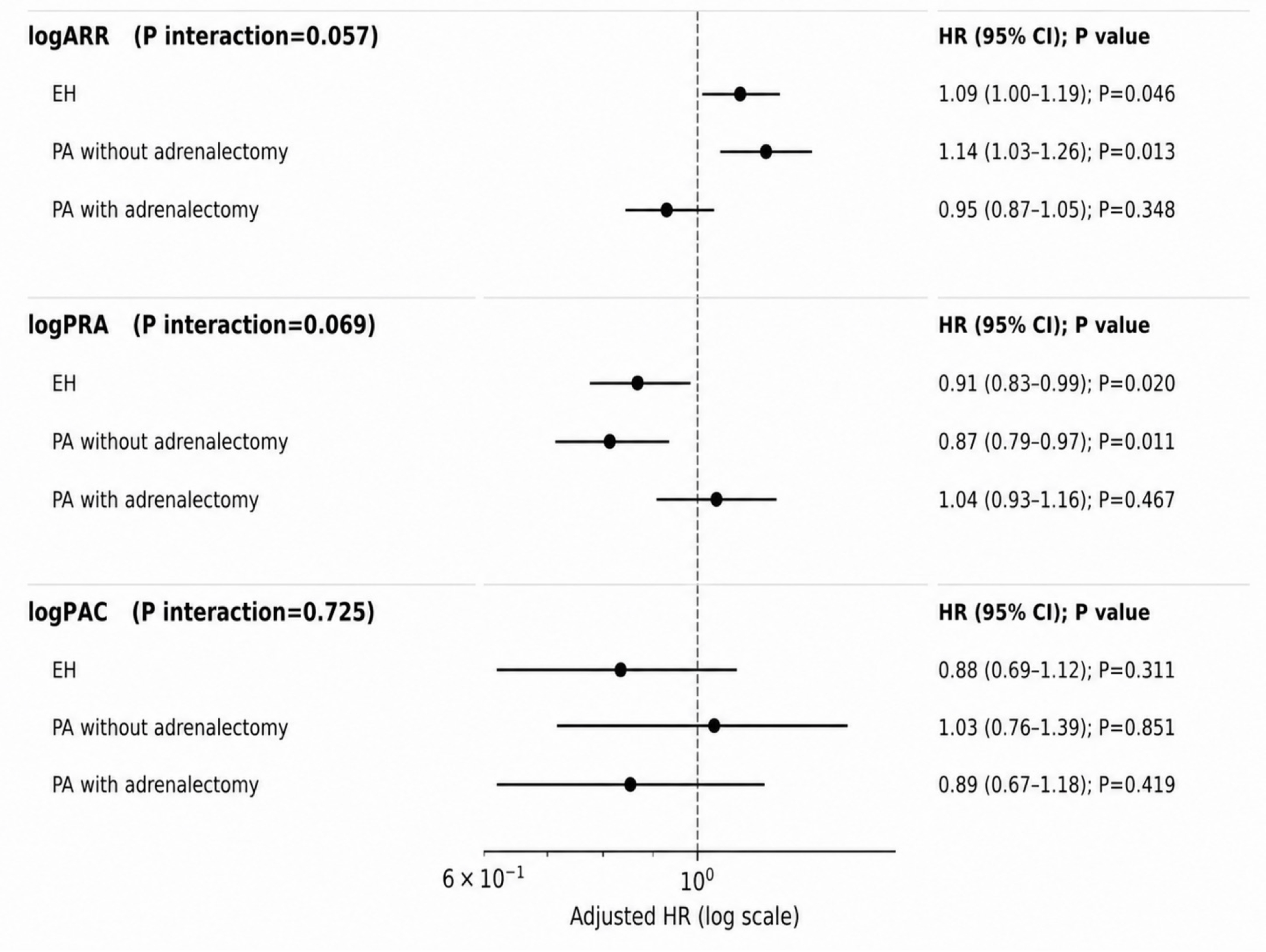
Associations of Screening Renin–Aldosterone Markers With Incident Major Adverse Cardiovascular Events. Adjusted hazard ratios and 95% confidence intervals are expressed per 1-unit increase in each natural-log-transformed biomarker and are shown separately for essential hypertension, primary aldosteronism without adrenalectomy, and primary aldosteronism with subsequent adrenalectomy. Each biomarker was analyzed in a separate model. Interaction P values test whether associations differ across clinical phenotypes. * Models were adjusted for prespecified age category (<65 vs ≥65 years), sex, body mass index, systolic blood pressure category (<150 vs ≥150 mmHg), estimated glomerular filtration rate, and serum potassium. *Abbreviations: ARR, aldosterone-to-renin ratio; EH, essential hypertension; MACE, major adverse cardiovascular events; PA, primary aldosteronism; PAC, plasma aldosterone concentration; and PRA, plasma renin activity.

**Table 3.** Adjusted Associations of Screening Renin–Aldosterone Indices With Incident MACE Across the Primary Aldosteronism Evaluation Pathway.

| **Exposure** | **EH** |  | **PA without OP** |  | **PA with OP** |  | **P for interaction** |
|---|---|---|---|---|---|---|---|
|  | **HR (95% CI)** | **P** | **HR (95% CI)** | **P** | **HR (95% CI)** | **P** |  |
| logARR | 1.09 (1.00–1.19) | 0.046 | 1.14 (1.03–1.26) | 0.013 | 0.95 (0.87–1.05) | 0.348 | 0.057 |
| logPRA | 0.91 (0.83–0.99) | 0.020 | 0.87 (0.79–0.97) | 0.011 | 1.04 (0.93–1.16) | 0.467 | 0.069 |
| logPAC | 0.88 (0.69–1.12) | 0.311 | 1.03 (0.76–1.39) | 0.851 | 0.89 (0.67–1.18) | 0.419 | 0.725 |

\* Hazard ratios are expressed per 1-unit increase in each natural-log-transformed biomarker. Each biomarker was analyzed in a separate model.
\* Models were adjusted for prespecified age category (<65 vs ≥65 years), sex, body mass index, systolic blood pressure category (<150 vs ≥150 mmHg), estimated glomerular filtration rate, and serum potassium.
\*Abbreviations: ARR, aldosterone-to-renin ratio; CI, confidence interval; EH, essential hypertension; HR, hazard ratio; OP, adrenalectomy; PA, primary aldosteronism; PAC, plasma aldosterone concentration; PRA, plasma renin activity.

Higher logPRA was associated with lower MACE risk in EH (adjusted HR, 0.91; 95% CI, 0.83-0.99; P=0.020) and PA without adrenalectomy (adjusted HR, 0.87; 95% CI, 0.79-0.97; P=0.011), but not in PA with adrenalectomy (adjusted HR, 1.04; 95% CI, 0.93-1.16; P=0.467). The logPRA-by-clinical-phenotype interaction did not reach statistical significance (P for interaction=0.069). Thus, ARR and PRA showed parallel qualitative patterns across the three clinical phenotypes.

LogPAC showed no consistent association with MACE across clinical phenotypes, with adjusted HRs of 0.88 (95% CI, 0.69-1.12; P=0.311), 1.03 (95% CI, 0.76-1.39; P=0.851), and 0.89 (95% CI, 0.67-1.18; P=0.419) in EH, PA without adrenalectomy, and PA with adrenalectomy, respectively. There was no evidence of effect modification by clinical phenotype for logPAC (p for interaction=0.725).

### Dose-Response Analyses

Cox generalized additive models showed broadly concordant patterns for ARR and PRA: MACE risk increased with higher logARR and decreased with higher logPRA in EH and PA without adrenalectomy, whereas both associations were relatively flat in PA with adrenalectomy (**eFigure 1**).

### Adrenalectomy Timing Sensitivity Analysis

In time-varying analyses among patients with PA, higher screening logARR was associated with MACE during the non-operated state (adjusted HR, 1.13; 95% CI, 1.03–1.25) but not during the operated state (adjusted HR, 0.95; 95% CI, 0.85–1.05; P for interaction=0.010). Similarly, higher screening logPRA was inversely associated with MACE during the non-operated state (adjusted HR, 0.88; 95% CI, 0.79–0.97) but not during the operated state (adjusted HR, 1.05; 95% CI, 0.94–1.18; P for interaction=0.016) (**eTable 2**). Operational post-treatment clinical and biochemical response classifications are summarized descriptively in **eTable 3**.

### Calendar-Era Sensitivity Analysis

After additional adjustment for index year, the effect estimates for logARR, logPRA, and logPAC were materially similar to those in the primary models. The biomarker-by-clinical-phenotype interaction reached statistical significance for logARR (P=0.022) and logPRA (P=0.033), but not for logPAC (P=0.825) (**eTable 4**). In era-stratified analyses, no significant biomarker-by-era interaction was observed within any clinical phenotype. Estimates in the 2020–2023 era were imprecise because of the limited number of MACE events, particularly among patients with PA who underwent adrenalectomy.

### Exploratory Negative Control Outcome Analyses

Exploratory analyses of abdominal hernia, lumbar/lumbosacral radiculopathy, gastrointestinal bleeding, and traffic accidents showed adjusted HRs of 0.837-1.193 for logARR and 0.834-1.156 for logPRA across clinical phenotypes (**eTable 5**).

## Discussion

In this structured PA evaluation cohort, higher screening ARR and lower screening PRA were associated with incident MACE among patients ultimately classified as EH or PA without adrenalectomy, whereas PAC alone showed no consistent association. Prior population studies linked low-renin, high-ARR phenotypes to cardiorenal events ^6,11–14^. Our findings indicate that higher ARR and lower PRA remain associated with MACE even after negative confirmatory testing for PA. Differences across treatment pathways require cautious interpretation because the primary biomarker-by-phenotype interactions were not statistically significant. In EH, negative confirmatory testing does not necessarily exclude risk-relevant renin–aldosterone physiology^8–10,32^.

In medically managed PA, the same ARR–PRA pattern may indicate persistent aldosterone-related risk and supports reassessment of MRA adequacy, renin recovery, potassium correction, and blood pressure control ^15,17,18,33,34^. Conversely, the absence of a clear association in the adrenalectomy pathway suggests that baseline screening physiology may be less informative after surgery, and post-treatment markers may warrant separate evaluation in future studies.

These findings show concordant associations of higher ARR and lower PRA with MACE, consistent with overlapping renin–aldosterone physiology. Prior population-based and hypertension cohorts have linked higher ARR and suppressed renin, but less consistently aldosterone concentration alone, with cardiovascular and kidney outcomes^11–14^. Our study extends this evidence to a clinically adjudicated PA pathway by distinguishing confirmatory-test-defined EH from overt PA and stratifying PA by subsequent adrenalectomy. This design evaluates whether screening ARR and PRA retain prognostic relevance after diagnostic classification and across treatment pathways.

### Comparative Interpretation of ARR and PRA

ARR and PRA provide different but overlapping views of renin–aldosterone physiology. Both measures showed qualitatively similar risk patterns in EH and PA without adrenalectomy, whereas PAC alone did not. This pattern accords with recent population-based evidence showing that both higher ARR and lower renin, but not aldosterone concentration alone, were associated with longitudinal kidney outcomes^13,14^. Our findings extend that comparative framework to MACE within a clinically defined PA evaluation pathway.

The EH finding deserves careful interpretation. Although these patients did not meet confirmatory criteria for PA, the EH group may include occult or subclinical PA phenotypes below conventional confirmatory thresholds^6,8–10^. Prior studies have linked higher ARR and suppressed renin to incident hypertension and cardiovascular events^8,11,12^. Long-term follow-up of patients with elevated ARR but initially negative confirmatory testing has also demonstrated progression toward more overt PA phenotypes^32^. In this cohort, higher ARR and lower PRA showed concordant associations with MACE after PA evaluation; these findings do not establish the added predictive value of using both markers together.

### Clinical Interpretation Across the PA Evaluation Pathway

Among patients with PA without adrenalectomy, higher ARR or lower PRA may each reflect a higher-risk renin-independent aldosterone phenotype. Persistent renin suppression in PA may indicate ongoing mineralocorticoid receptor activation and volume expansion despite blood pressure treatment, while ARR incorporates the relation between renin suppression and aldosterone concentration. This distinction is clinically relevant because outcomes in medically treated PA appear to depend not merely on whether MRA therapy is prescribed, but on the adequacy of mineralocorticoid receptor blockade, renin recovery, potassium correction, blood pressure control, and standardized medical response^15,17,18,33,34^. Thus, an abnormal ARR–PRA pattern in medically managed PA should prompt reassessment of treatment adequacy rather than be interpreted as a static diagnostic finding.

### Treatment Pathway, Adrenalectomy, and Residual Risk

In the main models, neither ARR nor PRA was associated with MACE among patients who underwent adrenalectomy. Targeted adrenalectomy can remove unilateral aldosterone excess, correct hypokalaemia, and improve blood pressure control^16,21^. Observational studies and meta-analyses have also reported favorable long-term cardiovascular outcomes after adrenalectomy in selected patients with PA^30,35,36^. However, treatment allocation was not randomized, and patients selected for adrenalectomy were likely to differ from non-operated patients^23,24^. Timing-sensitive analyses showed concordant attenuation of the ARR- and PRA-associated risk signals in the operated state. Confounding by indication and treatment-selection bias remain possible, and the exposure-by-clinical-phenotype interaction tests did not reach statistical significance.

The lack of a consistent association between PAC alone and MACE suggests that isolated aldosterone concentration may not capture the same risk-related physiology as ratio-based or renin-based measures. Aldosterone concentration varies with sodium intake, posture, potassium status, medications, and assay conditions^1,7^; ARR and PRA incorporate information about renin suppression in different ways^5,6^. Thus, the prognostic signal observed in this cohort appears to reflect renin– aldosterone imbalance rather than aldosterone concentration in isolation^13,14^.

### Clinical Implications

Our findings suggest a practical use for screening ARR and PRA after PA evaluation. In patients classified as EH, higher ARR and lower PRA were still associated with long-term MACE, indicating that negative confirmatory testing does not necessarily eliminate risk related to renin– aldosterone imbalance. These patients may merit closer cardiovascular risk assessment and repeat PA evaluation if clinical features evolve^1,32^. In PA without adrenalectomy, the same ARR–PRA pattern may indicate persistent aldosterone-related risk and may support reassessment of MRA adequacy, renin recovery, potassium correction, and blood pressure control^15,17,33,34^. In contrast, the lack of a clear association after adrenalectomy suggests that baseline screening physiology may be less prognostically informative in this pathway. Thus, higher ARR and lower PRA showed concordant associations with MACE after PA evaluation; their comparative and joint predictive utility, and their value in guiding treatment, remain to be established.

A recent analysis of the community-based Atherosclerosis Risk in Communities cohort associated higher ARR with incident atrial fibrillation and ischemic stroke^37^. Unlike that study, our cohort comprised patients undergoing structured PA evaluation, allowing prognostic associations to be examined after confirmatory diagnostic classification and across subsequent management pathways. Whether these associations improve cardiovascular risk prediction beyond conventional clinical factors remains to be established.

### Strengths and Limitations

Strengths of this study include evaluation of screening ARR, PRA, and PAC within a structured PA diagnostic pathway, confirmatory-test-based clinical classification, linkage to long-term nationwide outcomes, and assessment across subsequent treatment pathways. This design allowed screening renin–aldosterone physiology in hypertensive patients to be examined in a clinically adjudicated setting rather than only in general population-based cohorts. Several limitations should be acknowledged. First, this cohort consisted of patients selected for PA evaluation and should not be interpreted as representative of an unselected hypertension population. Referral for PA testing reflected resistant hypertension, hypokalemia, adrenal abnormalities, or other clinical concerns that were not fully captured in the available data. Second, to reduce medication-related interference with ARR, PRA, and PAC, potentially interfering antihypertensive agents were withheld for at least 21 days when clinically appropriate, with doxazosin or diltiazem used for blood pressure control; usual sodium intake was maintained and assessed by urinary testing. Third, ARR and PRA are mathematically coupled because PRA is the denominator of ARR; therefore, their concordant associations should be interpreted in the context of overlapping physiology rather than as statistically independent effects. Separate models allow the associations of ARR and PRA to be evaluated under the same covariate framework but do not establish biological independence. Fourth, the cohort spanned a prolonged period during which PA screening, confirmatory testing, subtype evaluation, and treatment allocation may have evolved. Although additional adjustment for index year and era-stratified analyses showed no significant biomarker-by-era interaction, estimates in the most recent era were imprecise because of sparse events; temporal heterogeneity therefore cannot be completely excluded. Fifth, adrenalectomy was a post-baseline, nonrandomly assigned treatment pathway. Moreover, in the primary phenotype-stratified models, the exposure-by-clinical-phenotype interaction tests for logARR and logPRA did not reach statistical significance. Finally, exploratory negative control analyses showed no consistent associations with screening logARR or logPRA. However, some estimates were imprecise, and these analyses cannot exclude residual confounding.

### Perspectives

These results indicate that a negative confirmatory assessment for PA does not necessarily eliminate the cardiovascular prognostic information carried by screening ARR and PRA among patients selected for PA evaluation. Renin–aldosterone measures could provide physiological context for cardiovascular risk assessment after diagnostic workup, particularly for patients managed without adrenalectomy. However, ARR and PRA share the same renin component, and their concordant associations do not demonstrate independent predictive gains. Treatment-pathway differences are vulnerable to confounding and selection; the primary interactions were not statistically significant. Further studies should test whether serial renin assessment, adequacy of PA treatment, and validated risk prediction models improve patient management.

## Novelty and Relevance

### What Is New?

- In a seven-hospital PA evaluation cohort linked to nationwide outcomes, screening ARR, PRA, and PAC measured before confirmatory testing were evaluated in parallel for their associations with long-term MACE.
- Associations were examined across confirmed EH, medically managed PA, and subsequent adrenalectomy pathways.

### What Is Relevant?

- Higher screening ARR and lower PRA remained associated with MACE even in patients without confirmed PA.

### Clinical/Pathophysiological Implications?

- Renin–aldosterone physiology may inform cardiovascular risk assessment after PA workup.

## Data Availability

Individual-level data from the Taiwan Primary Aldosteronism Investigation (TAIPAI) registry linked to the National Health Insurance Research Database (NHIRD) are not publicly available because of privacy and regulatory restrictions. Aggregated results and operational definitions are provided in the manuscript and supplemental materials.

## Data Sharing

Individual-level data are not publicly available because of privacy and regulatory restrictions governing the NHIRD and institutional registry data.

## Acknowledgments

The authors thank the staff of the Community Medicine Research Center of Keelung Chang Gung Memorial Hospital. We also express our sincere gratitude to the administrative support of the Chang Gung Memorial Hospital Clinical Trial Center and all staff of the Taiwan Clinical Trial Consortium (TCTC). VCW had full access to all the data in the study and takes responsibility for the integrity of the data and the accuracy of the data analysis.

## Use of Artificial Intelligence in Manuscript Preparation

ChatGPT (OpenAI) was used during manuscript preparation and revision in 2026, including revisions on November 5, 2026, to assist with English-language editing, manuscript organization, literature review and synthesis, and journal-specific formatting. No extensions were used. Python code developed with assistance from ChatGPT was used to recreate and format Figures 1 and 2 from author-provided results. The exact model versions used in earlier manuscript-preparation and figure-formatting sessions could not be retrospectively verified. AI was not used to generate or modify the underlying research data or to perform the primary statistical analyses. All AI-assisted content, including manuscript text, cited references, figures, and statistical values, was reviewed and verified by the authors, who take full responsibility for its accuracy and integrity.

## Contributors

All authors contributed significantly to the design and conduct of the study and acquisition of clinical data. HCP and VCW conceived the study. HCP conducted data analyses and wrote the first draft of the manuscript. KHC supervised all analyses, assumes responsibility for the analyses, and assumes responsibility for data interpretation. CHT and YHL reviewed and commented on the manuscript. CYW and VCW constructed the dataset, including the biomarker measurements in each participant, and reviewed and commented on the drafts of the manuscript.

## Supplemental Material

Supplemental Methods

eTables 1–5

eFigure 1

